# Epigenetic Signatures Reveal Biological Embedding of the Early-Life Environment Two Decades after Exposure to Adversity

**DOI:** 10.64898/2026.01.28.26345020

**Authors:** Archibold Mposhi, Megan Buchanan, Sophie B. Mériaux, Jeanne Le Cleac’H, Martha M.C. Elwenspoek, Fleur A.D. Leenen, Claude P. Muller, Claus Vögele, Jonathan D. Turner

**Affiliations:** Department of Infection and Immunity, Luxembourg Institute of Health, Esch-sur-Alzette, Luxembourg; Faculty of Science Technology and Medicine, University of Luxembourg, Belval Luxembourg; Department of Behavioural and Cognitive Sciences, University of Luxembourg, Belval, Luxembourg

**Keywords:** DNA methylation, early-life adversity, HPA axis, institutionalisation, health trajectories

## Abstract

**Introduction:** Early-life adversity (ELA) encompasses a range of environmental stressors, including physical, emotional, and social challenges that can affect health during the critical early developmental period. Extensive research has linked ELA to negative long-term health outcomes, yet the underlying biological mechanisms remain poorly understood. The current study investigates how early institutional care changes the epigenetic landscape in young adults. The study also provides insights into role of DNA methylation as a potential mediator for disease susceptibility and altered health trajectories.

**Materials and Methods:** DNA was extracted from blood samples obtained from 111 individuals (71 Controls; 40 ELA) who were part of the EpiPath cohort. DNA methylation was measured using the Infinium Methylation EPIC v2.0 BeadChip.

**Results:** 3,785 differentially methylated CpG loci were identified in the ELA group in comparison with the control group (FDR <0.05). Pathway enrichment analysis highlighted biological processes involved in metabolic regulation, stress response, and neurodevelopment, with novel pathways such as GTPase-mediated signalling, efferocytosis and glucuronosyltransferase emerging as potential drivers of the ELA phenotype. A subset of 28 CpG loci was used to develop an epigenetic signature, which showed a significant association with the development of chronic diseases in ELA-exposed individuals.

**Conclusion:** This study reinforces Barker’s concept of sensitive periods and it underscores the enduring impact of ELA in shaping long-term health outcomes. The persistence of DNA methylation patterns decades after exposure to ELA, and their clear association with the resultant phenotype confirms that stable epigenetic imprints play a potential role in long-term disease risk and resilience.

## 1. Introduction

The study of early-life adversity (ELA) has highlighted the significance of an individual’s first 1,000 days of life in relation to adulthood health and disease [1]. ELA refers to environmental stressors involving physical, emotional, financial and social aspects that influence both health and development starting from conception [2]. The Development of Health and Disease (DOHaD) or the “Barker theory” has given weight to the importance of sensitive periods, particularly early life, where individuals in adverse conditions risk becoming more susceptible to disease in later life [3]. The early-life developmental window highlights the significance of timing for adverse stressors where many biological systems may potentially adjust and set themselves to the experienced environment [4]. This follows the “three-hit” model by Daskalakis et al. in 2013 where genetic factors from conception interact with the early-life exposome to produce latent phenotypes that may increase susceptibility to certain diseases in late-life environments [5]. Significant gaps remain in our understanding of the specific biological pathways that connect ELA to long-term health outcomes, particularly the interplay between stress reactivity, immune dysregulation, and epigenetic modifications.

Adult health issues involving cardiovascular disease [6] and metabolic diseases like type 2 diabetes mellitus [7] have been linked to these negative early exposures. Certain psychopathologies such as depression and post-traumatic stress disorder have also been associated with exposure to early trauma that may be related to changes in genes associated with stress regulation [8]. Researchers have noted other key findings concerning epigenetic changes during this sensitive window, including lifelong effects on stress regulation for mothers and infants possibly due to the interaction between the hypothalamic-pituitary-adrenal (HPA) axis and oxytocinergic systems during postpartum skin-to-skin contact and breastfeeding [9]. The importance of the mother-infant bond is underscored by research linking early parental separation and institutionalisation or adoption in childhood with negative health outcomes [10]. Overall, institutional care with non-continuous carers has been argued as one of the worst areas of privation and neglect for children and developmental assessments have shown delays in both cognitive and motor functions [11]. Institutionalisation in early age, particularly during infancy, has also been shown to promote higher levels of anxiety [12] and amygdala activity associated to perceived threats [13] that may continue after adoption within nurturing familial environments. Furthermore, epigenetic consequences may be linked to institutionalisation, particularly with changes in gene expression due to alterations in DNA methylation at certain CpG sites that potentially reprogram immune system functioning due to modifications such as lower B-cells and T-helpers [14].

Despite the increased susceptibility to disease and psychopathology, adolescence has been identified as a potential period for modifying the HPA axis towards healthier functioning by altering personal interpretations of environmental stressors [15]. These findings highlight the benefits of preventive measures towards identifying individuals of ELA backgrounds who are at greater risk for developing adverse physical and psychological conditions. This study utilised data from participants of the EpiPath cohort comprised of individuals who had experienced ELA operationalised as early parental separation with the majority being institutionalised at birth [16]. This cohort is uniquely suited to examine the long-term impacts of severe ELA on stress reactivity and immune function. Despite the potential recalibration period during adolescence, previous findings from this cohort revealed that ELA individuals exhibited inhibited stress reactivity within the HPA axis [17] and elevated levels of proinflammatory T cells such as Th17 [18] even into early adulthood. In addition to these findings, Naumova et al. (2012) observed increased methylation in genes largely associated with immune and cellular signalling functions in children who had been institutionalised [11]. These findings suggest that ELA may exert lasting biological effects that are potentially mediated by epigenetic modifications and immune dysregulation.

Through this study, we aim to address critical gaps in knowledge on the mechanisms underlying ELA mediated health outcomes. Here, we explore how adverse early-life environments such as institutionalisation alter the epigenetic landscape of exposed children and we highlight how this may persist to shape their health trajectories.

## 2. Materials and Methods

### 2.1 Human Cohort

The study used samples and data from our previously published EpiPath cohort. EpiPath is a cohort of 115 young adults recruited from July 2014 until March 2016 within Luxembourg and the greater region [18]. The control group was comprised of 73 individuals who were raised by their biological parents. The remaining 42 participants underwent parental separation and adoption at an early age with the majority also experiencing institutional care (Figure 1). For those formerly institutionalised, the median adoption timeframe was 4 months [19]. Our cohort for this study consisted of 111 participants from the EpiPath study for whom DNA samples remained.

**Figure 1:**
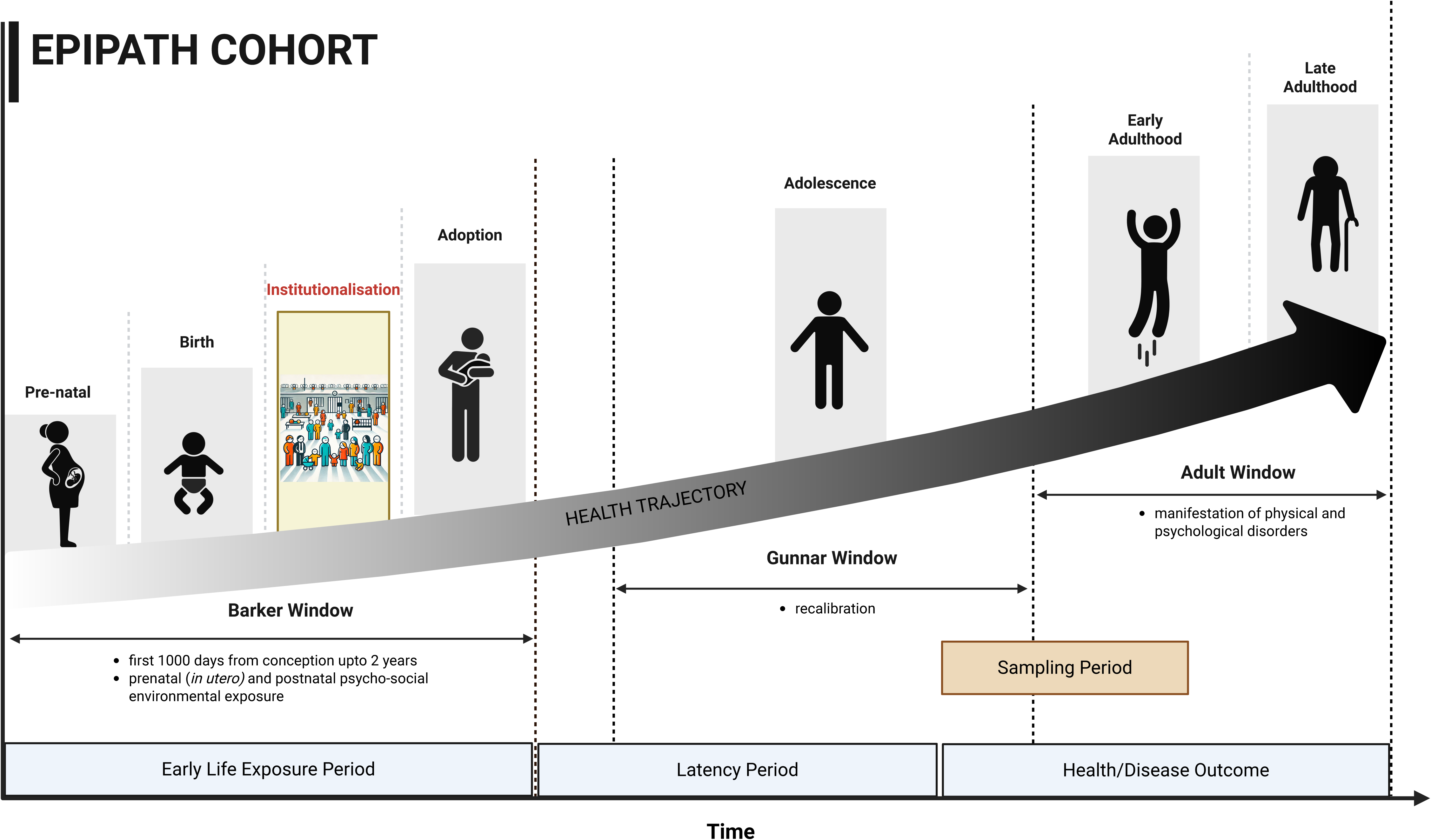
Graphical representation of the EpiPath Cohort and time-line. Illustration shows the different windows of development from early life to late adulthood. The Baker window denotes the early life period from conception right up to the age of 2 years. Here individuals are exposed to ELA in the form of institutionalisation and then adopted at a median age of 4.3 months. This is followed by the Gunnar window, where individuals undergo potential recalibration, which can either propel or change the overall trajectory and subsequent health outcome. Blood samples from EpiPath participants were collected during the late pubertal to early adulthood period.

The EpiPath study received ethical approval by the National Research Ethics Committee of Luxembourg (CNER, reference 201303/10 v1.4) and the University of Luxembourg Ethics Review Panel (ERP, No 13–002). The current study was performed in accordance to guidelines from the revised 2013 Declaration of Helsinki and the Directive 2001/20/EC of the European Union. All participants gave their written informed consent in addition to receiving a small financial compensation for inconvenience and time.

### 2.2 DNA Extraction

All blood samples were collected at a fixed time (11 o’clock in the morning) before being sent for immediate processing. Automated DNA extraction was performed on the QIAcube (Qiagen, Venlo, Netherlands) on 200μl of whole blood using the QIAamp DNA Mini kit (Qiagen). Subsequently, 500ng of DNA was bisulfite-converted using the EZ DNA Methylation™ Kit (Zymo, Freiburg, Germany) according to the manufacturer’s instructions.

### 2.3 DNA Methylation Analysis

This studied analysed the DNA methylation patterns using the Infinium Methylation EPIC v2.0 BeadChip (Illumina, Eindhoven, The Netherlands) and iScan (Illumina). The Infinium HD Methylation Assay Reference Guide was followed for all processing of samples alongside the GenomeStudio Software 2.0 (Illumina) for quality assessment.

Briefly, the 111 DNA samples underwent bisulfite conversion with the EZ DNA Methylation Kit (Zymo Research, Irvine, California, US) and were stored at −20°C until use. Following the manufacturer’s recommendations for whole-genome amplification, the bisulfite-converted DNA was denatured and isothermally amplified at 37°C in a hybridization oven for 24 hours using the kit from Illumina. The amplified DNA samples went through an endpoint enzymatic fragmentation followed by the isopropanol precipitation and resuspension processes. Samples were hybridized to the Infinium microarray probes overnight at 48°C. The BeadChips were washed with the provided buffer before being placed in a temperature-controlled chamber at 44°C for the single base extension reaction to label probes. Afterwards beadchips were incubated at 32°C for the fluorescent staining process. Lastly, the Illumina iScan platform was used to scan the dried sample arrays for imaging of CpG loci with default settings.

### 2.4 Data Analysis

For preliminary quality control assessment data were exported as .idat files and uploaded into GenomeStudio (Illumina). The .idat files were also imported into R Studio (R version 4.4.1) and data analysis was performed using Illumina’s recommended SeSAMe pipeline (Sensible step-wise analysis of DNA methylation bead chips) for quality pre-processing of data. SeSAMe R package version 1.22.2 was used [20]. The raw IDAT files were read into the R workspace as sigDF objects using the readIDATpair() function with the platform parameter set to “EPICv2”. Data pre-processing was performed with the openSesame() function, using the recommended human-specific preparation code (“QCDPB”) as the prep argument. Probes with detection p-values greater than 0.05 were filtered out, out-of-band probes were used for signal background correction, and quantile normalization was applied.

To address batch effects, the sva R package (version 3.52.0; [21]) combat() function was used. Next we excluded a total of 120,937 probes associated with single nucleotide polymorphisms (SNPs) from the analysis to minimize DNA methylation bias. Additionally, probes mapping to the X and Y chromosomes were filtered out. A matrix of β-values was generated using the getBetas() function, annotated with sample metadata and the manifest file corresponding to the hg38 genome build (https://zwdzwd.github.io/InfiniumAnnotation#human/).

Differential methylation analysis was performed using sesame (DML() function) with the sample group specified as the contrast. In the analysis, age, smoking, sex, body mass index (BMI) and Cytomegalovirus (CMV) infection status were included as covariates. A summary table of the results was obtained using the summaryExtractTest() function and results with an FDR < 0.05 were identified. Partial least squares-discriminant analysis (PLS-DA) was performed using the mixOmics R package (version 6.28.0) and hierarchical clustering, gene ontology, KEGG pathway and reactome enrichment analysis of data was performed in R using clusterProfiler (version 4.12.6). Differentially methylated regions and transcription factor binding sites were analysed using sesame’s DMR() function and testEnrichment() function respectively. PCs were extracted and associated using cor.test() function in the general R stats package (version 4.4.1). Mediation models were performed in R using mediation (version 4.5.0). Data figures were generated using ggplot2 (version 3.5.1 [22]), cowplot (version 1.1.3; [23]), corrplot (version 0.94; [24]), ComplexHeatmap (version 2.20.0; [25]) and pheatmap (version 1.0.12; [26]).

## 3. Results

### 3.1. Cohort Overview

The final analysis was performed on 71 controls and 40 ELA participants. The control and ELA group participants were matched for sex (p=0.354, χ^2^ test), but did show a small significant difference in age (control: median, 22y [IQR, 21-23]; ELA: median, 24y [IQR, 20-25.5]; p=0.036, Wilcoxon rank-sum test). The ELA group underwent adoption at the median age of 4.3 months (Table 1). As adoption occurred at a young age, the CTQ scores which highlight memories of traumatic experiences before age 16 reflected similar scores between both groups.

**Table 1:** Cohort Overview and Demographics.

|  | *Variable* | *Controls (n = 71)*<br>*(Median, IQR)* | *ELA (n = 40)*<br>*(median, IQR)* |
| --- | --- | --- | --- |
| *Demographics* | Age (years) | 22 (21-23) | **24.0 (20.0-25.5)\*** |
|  | Sex (women) | 57% | 68% |
| *Health* | Chronic disease | 9% | **43%\*\*\*** |
| *ELA immune phenotype* |  |  |  |
| *Immune activation* | Plasma CRP (mg/L) | 2.0 (1.3-3.3) | 2.0 (1.1-3.9) |
|  | Plasma IL-6 (pg/mL) | 0.6 (0.3-1.1) | 0.7 (0.3-1.1) |
|  | IL-6 secretion (sum z-score) | 0.7 (-0.7-1.7) | **-0.4 (-2.6-1.2)\*** |
| *Cellular immunity* | HSV-1 seropositive/titers<sup>1</sup> | 37% / 1.9 (1.7-2.0) | 45% / 1.8 (1.7-2.0) |
|  | EBV seropositive/titers<sup>1</sup> | 46% / 1.9 (1.4-2.3) | **68%<sup>#</sup> / 2.1 (1.5-2.7)** |
|  | CMV seropositive/titers<sup>1</sup> | 12% / 1.3 (1.1-1.9) | 19% / 1.2 (1.1-1.2) |
| *Immunosenescence* | Telomere length (relative T/S) | 1.2 (0.8/1.9) | 1.1 (0.7-1.7) |
| *Stress* | Salivary cortisol (µg/dL) | 0.26 (0.17-0.34) | 0.23 (0.19-0.34) |
|  | Stress rating (Visit 1) | 1.2 (0.6-2.6) | 1.5 (0.2-3.1) |
| *Health behaviours* | Smoking | 12% | **30%\*** |
|  | BMI (kg/m<sup>2</sup>) | 22.6 (20.2-24.3) | **23.2 (21.3-26.8)\*** |
|  | Physical activity (IPAQ, METs) | 3772 (1706-5784) | 3051 (1452-8633) |
| *Adoption* | Age (months) | 0 (0-0) | 4.3 (0-15) |
| *Trauma* | Childhood trauma (CTQ) | 1.2 (1.1-1.4) | 1.2 (1.1-1.4) |

<sup>1</sup>Among seropositive participants.
<sup>#</sup>p<0.10, \*p<0.05, \*\*\*p<0.001, compared with control group.
Statistical analyses performed using a Wilcoxon rank-sum test for numerical values and Pearson's chi-square test for categorical variables.
Abbreviations: IQR, interquartile range; CRP, C-reactive protein; IL-6, interleukin 6; HSV-1, Herpes simplex 1; EBV, Epstein-Barr virus; CMV, Cytomegalovirus; BMI, Body mass index; IPAQ, international physical activity questionnaire; METs, metabolic equivalent of task units; T/S, ratio of telomere and single copy gene (36B4) cycle threshold values; CTQ, childhood trauma questionnaire.

### 3.2. Differential DNA methylation in individuals exposed to early life institutional care

In order to identify the differentially methylated CpG loci an epigenome-wide-association study (EWAS) was conducted. A total of 3,785 differentially methylated CpG loci were identified in the ELA group following false discovery rate (FDR) correction for multiple testing (FDR < 0.05; Figure 2A). Of these, a decrease in DNA methylation was observed in 2139 CpG loci associated with 1397 genes while an increase in DNA methylation was seen in the remaining 1646 CpG loci associated with 1012 genes (FDR < 0.05) (Figure 2B – C, Supplementary Table 1). To determine whether ELA contributes to the observed variability in DNA methylation profiles between the control group and ELA exposed group, we conducted a Principal Component Analysis (PCA). The first principal component (PC1) accounted for 24.69% of the total variance, while the second principal component (PC2) explained 5.57% of the variance (Figure 2D).

**Figure 2.**
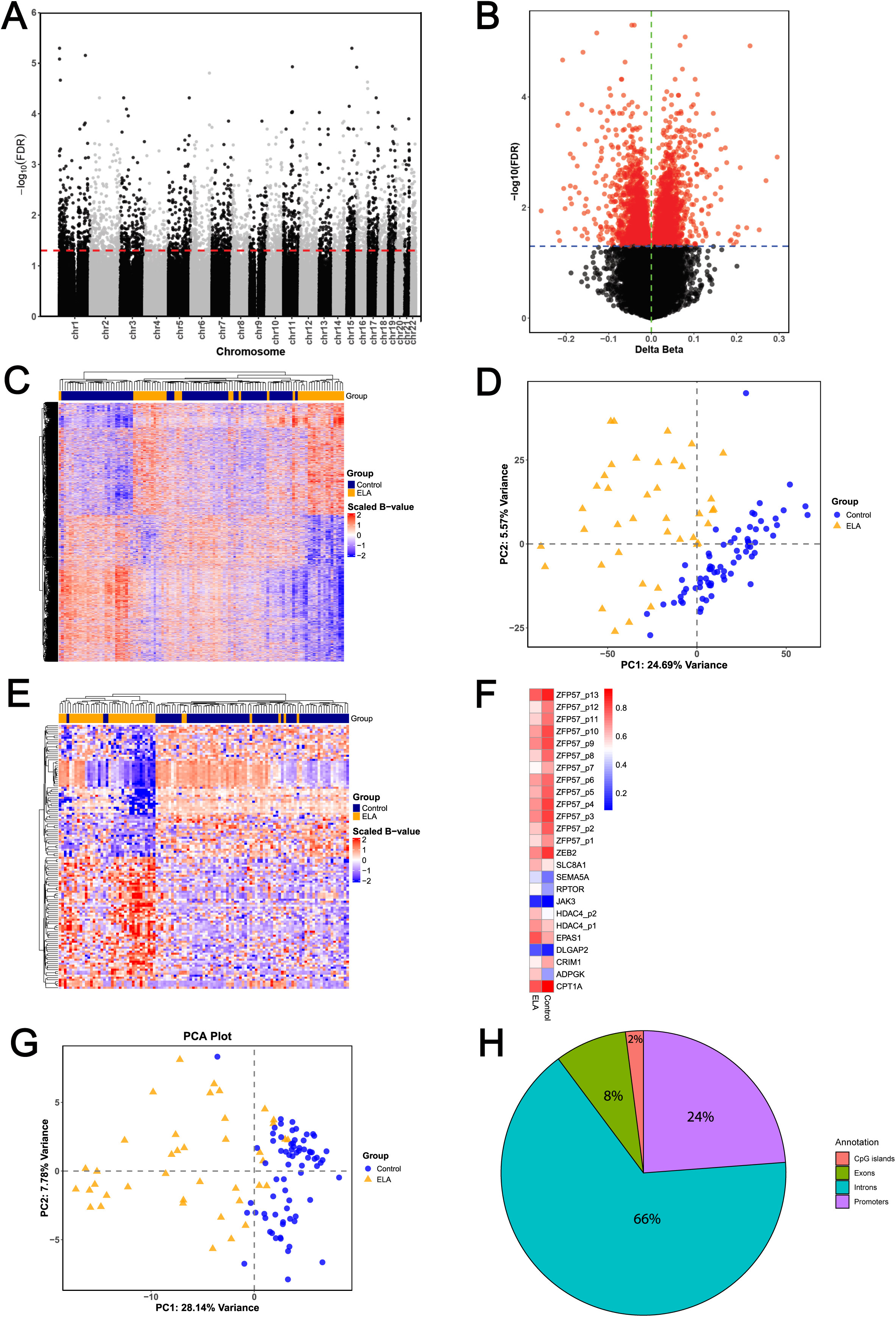
Differential DNA methylation analysis of individual CpG loci in ELA-exposed group. **(A)** Manhattan plot shows genome-wide differential methylation of 3785 CpG loci identified in the ELA group. Red line depicts the FDR correction threshold (FDR < 0.05). **(B)** Volcano plot illustrates the distribution of the 3785 differentially methylated CpG loci. Blue line represents the significance threshold and red dots represent all significant probes (FDR < 0.05). **(C)** Heatmap shows DNA methylation levels of the 3785 CpG loci in the ELA and control groups (FDR < 0.05). **(D)** Principal Component Analysis (PCA) plot based on all differentially methylated CpG loci. **(E)** Heatmap of the 132 most biologically relevant differentially methylated CpG loci (FDR < 0.05, effect size > 0.1), highlighting methylation differences between ELA and control groups. **(F)** Heatmap of mean DNA methylation levels for CpGs associated with key genes implicated in stress response, neurodevelopment, immune response, mitochondrial metabolism, and epigenetic imprinting. **(G)** PCA plot based on the 132 most biologically relevant CpG loci. **(H)** Pie chart depicting the genomic distribution of the 132 CpG loci across CpG islands, promoter regions, exons, and introns.

To identify the most biologically relevant differentially methylated CpG loci, we applied a stringent selection criterion: FDR < 0.05 and an effect size > 0.1. Using this approach, 132 CpG loci associated with 80 genes, encompassing both protein-coding genes and long non-coding RNAs were identified (Figure 2E; Supplementary Table 2). Of these, 66 CpG loci linked to 33 genes showed a decrease in DNA methylation, while the other 66 CpG loci, associated with 47 genes, showed increased DNA methylation. Of note, these genes were associated with stress response and neurodevelopment (DLGAP2, EPAS1, HDAC4, ZEB2), immune response (CRIM1, JAK3, SEMA5A), mitochondrial metabolism (ADPGK, CPT1A, SLC8A1) and epigenetic imprinting (ZFP57) (Figure 2F).

Next we conducted a PCA on the 132 CpG loci. PC1 accounted for 28.14% of the total variance, while PC2 explained 7.78% of the variance (Figure 2G). In order to increase the depth of our analysis we explored the genomic context of the 132 differentially methylated CpGs. 2.04% of the CpGs were located on CpG islands while 23.81% were found on promoter regions. In addition, only 8.16% of the probes were located in exonic regions while 65,99% were within introns (Figure 2H).

### 3.3. DNA methylation within DMRs reveals potential epigenetic modulation of gene expression

To expand our analysis beyond individual CpG loci, we investigated differentially methylated genomic regions (DMRs). This approach aggregates neighbouring CpGs with consistent differential methylation into genomic location-based DNA segments. DMRs were filtered to include only those with an adjusted FDR p-value < 0.05. Using this method, we identified 8,417 DMRs spanning all 22 chromosomes in the ELA group compared to the control group (Figure 3A-C). Within these regions, we found CpGs associated with 3,488 protein-coding genes, 1,292 long non-coding RNAs (lncRNAs), 10 microRNAs (miRNAs), 4 small nucleolar RNAs (snoRNAs), 4 miscellaneous RNAs (miscRNAs), and 3 small nuclear RNAs (snRNAs) (Supplementary Table 3).

**Figure 3:**
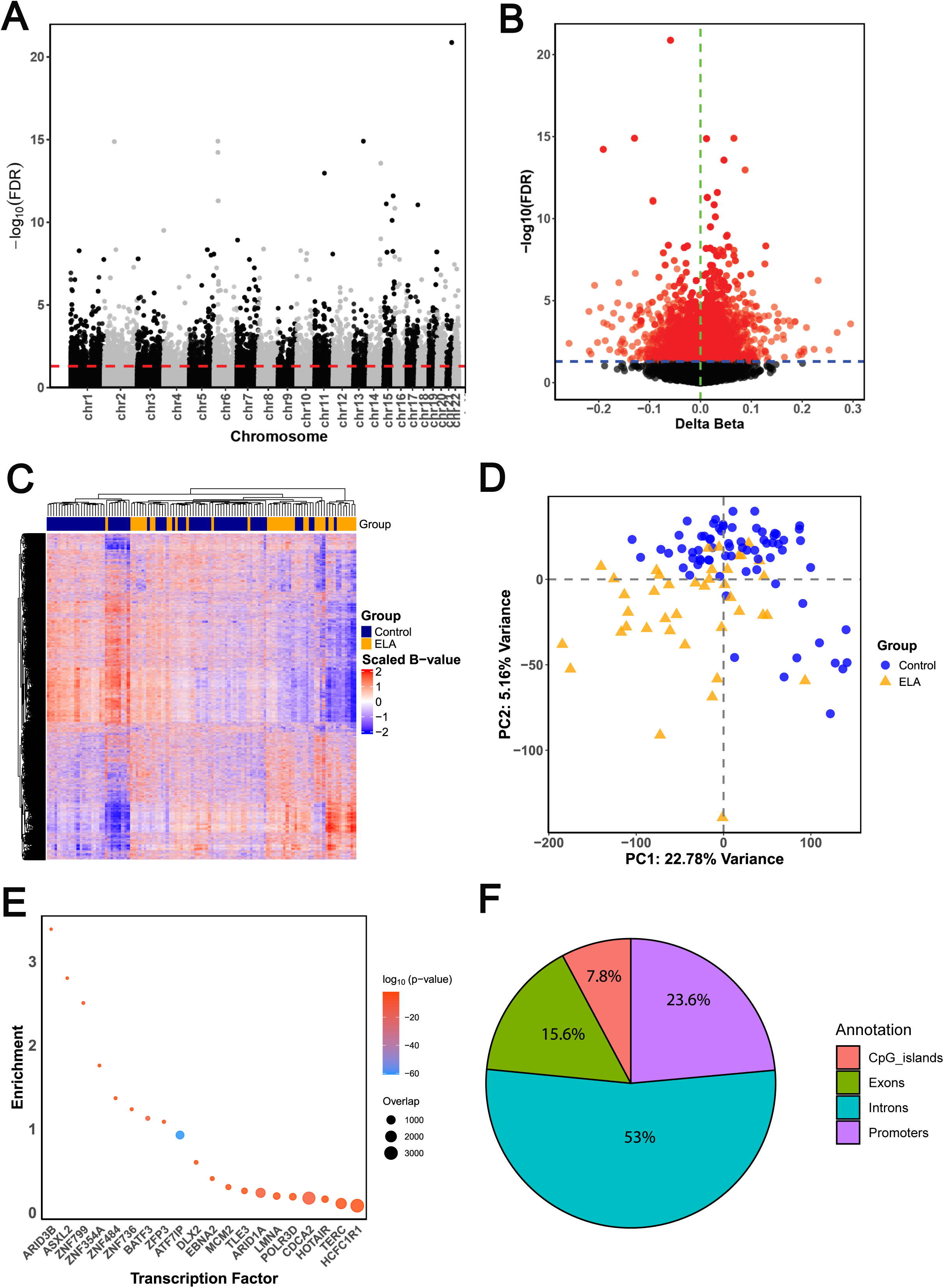
DNA methylation analysis of differentially methylated regions (DMRs) and transcription factor binding enrichment analysis. **(A)** Manhattan plot shows genome-wide differential methylation of CpG loci within 8 417 significant DMRs. The red dotted line represents the significance threshold (FDR <0.05). **(B)** Volcano plot shows the distribution of CpG loci within the 8 417 DMRs. The blue dotted line depicts the significance threshold (FDR <0.05) and all red dots represent significant loci. **(C)** Heatmap shows DNA methylation for all CpG loci within the significant DMRs. **(D)** Principle Component Analysis (PCA) plot based on all significant DMRs. **(E)** Transcription factor binding enrichment analysis based on all the significant DMRs. **(F)** Pie chart depicting the genomic distribution of the DMRs across CpG islands, promoter regions, exons, and introns.

Furthermore, a PCA of the DMRs showed that PC1 accounted for 22.78% of the total variance while PC2 explained 5.16% of the variance (Figure 3D). Next, we performed transcription factor binding site enrichment analysis (TFBS) to identify the transcription factors that were enriched in the DMRs. We identified 5 significantly enriched transcription factor-binding sites in the DMRs (FDR < 0.05)(Table 2, Figure 3E). We observed a significant enrichment of ATF7IP, BATF3, CDCA2, ARID1A and ZNF799 binding sites in the DMRs (FDR< 0.05, F). In order to define the genomic context of the DMRs, we annotated the CpGs within these regions. 7.8% of the CpGs were located on CpG islands while 23.6% were found on promoter regions. In addition, 15.6% of the probes were located in exonic regions while 53% were within introns (Figure 3F). These results suggest that ELA-mediated DNA methylation may affect a myriad of pathways involved in the regulation of gene expression, with potential implications for transcriptional regulation and cellular function.

**Table 2:** Transcription Factor Binding Site Analysis (TFBS).

| *Transcription Factor* | *Enrichment* | *Overlap* | *P.value* | *FDR* |
| --- | --- | --- | --- | --- |
| *ATF7IP* | 0.93 | 858 | 1.53E-61 | 2.07E-58 |
| *BATF3* | 1.13 | 77 | 1.50E-09 | 1.02E-06 |
| *CDCA2* | 0.17 | 2689 | 1.02E-08 | 4.59E-06 |
| *ARID1A* | 0.24 | 1316 | 1.67E-08 | 5.66E-06 |
| *ZNF799* | 2.50 | 10 | 2.51E-05 | 6.80E-03 |
| *ZFP3* | 1.09 | 24 | 8.02E-04 | 1.81E-01 |
| *TLE3* | 0.26 | 310 | 1.37E-03 | 2.55E-01 |
| *LMNA* | 0.20 | 522 | 1.50E-03 | 2.55E-01 |
| *TERC* | 0.11 | 1780 | 1.73E-03 | 2.60E-01 |
| *POLR3D* | 0.19 | 530 | 2.02E-03 | 2.60E-01 |
| *MCM2* | 0.30 | 207 | 2.11E-03 | 2.60E-01 |
| *HCFC1R1* | 0.08 | 3055 | 2.34E-03 | 2.65E-01 |
| *DLX2* | 0.60 | 55 | 2.56E-03 | 2.67E-01 |
| *ZNF736* | 1.23 | 15 | 2.85E-03 | 2.76E-01 |
| *ZNF484* | 1.37 | 12 | 3.61E-03 | 3.17E-01 |
| *ASXL2* | 2.80 | 4 | 3.74E-03 | 3.17E-01 |
| *ARID3B* | 3.39 | 3 | 4.47E-03 | 3.57E-01 |
| *ZNF354A* | 1.76 | 7 | 6.33E-03 | 4.77E-01 |
| *HOTAIR* | 0.16 | 489 | 9.65E-03 | 6.89E-01 |
| *EBNA2* | 0.41 | 75 | 1.19E-02 | 7.74E-01 |

### 3.4. Adversity during early life leaves an epigenetic imprint several years after initial exposure

To identify an epigenetic signature capable of distinguishing the ELA group from the control group, we performed a Partial Least Squares Discriminant Analysis (PLS-DA). This supervised method combines dimensionality reduction with the predefined classifications. Component 1 accounted for 10% of the total variance, while Component 2 explained 7% of the variance (Figure 4A). To evaluate the classification accuracy of the PLS-DA model, we performed a cross-validation. In order to refine the model’s classification power and to avoid overfitting the model, we included Component 3, which accounted for 4% of the total variance. Using three components, the overall classification error and balanced error rate (BER) based on Mahalanobis distance was 0.22 ± 0.08 and 0.27 ± 0.07, respectively, demonstrating an improved model performance compared to a single-component classification. (Figure 4B – C).

**Figure 4:**
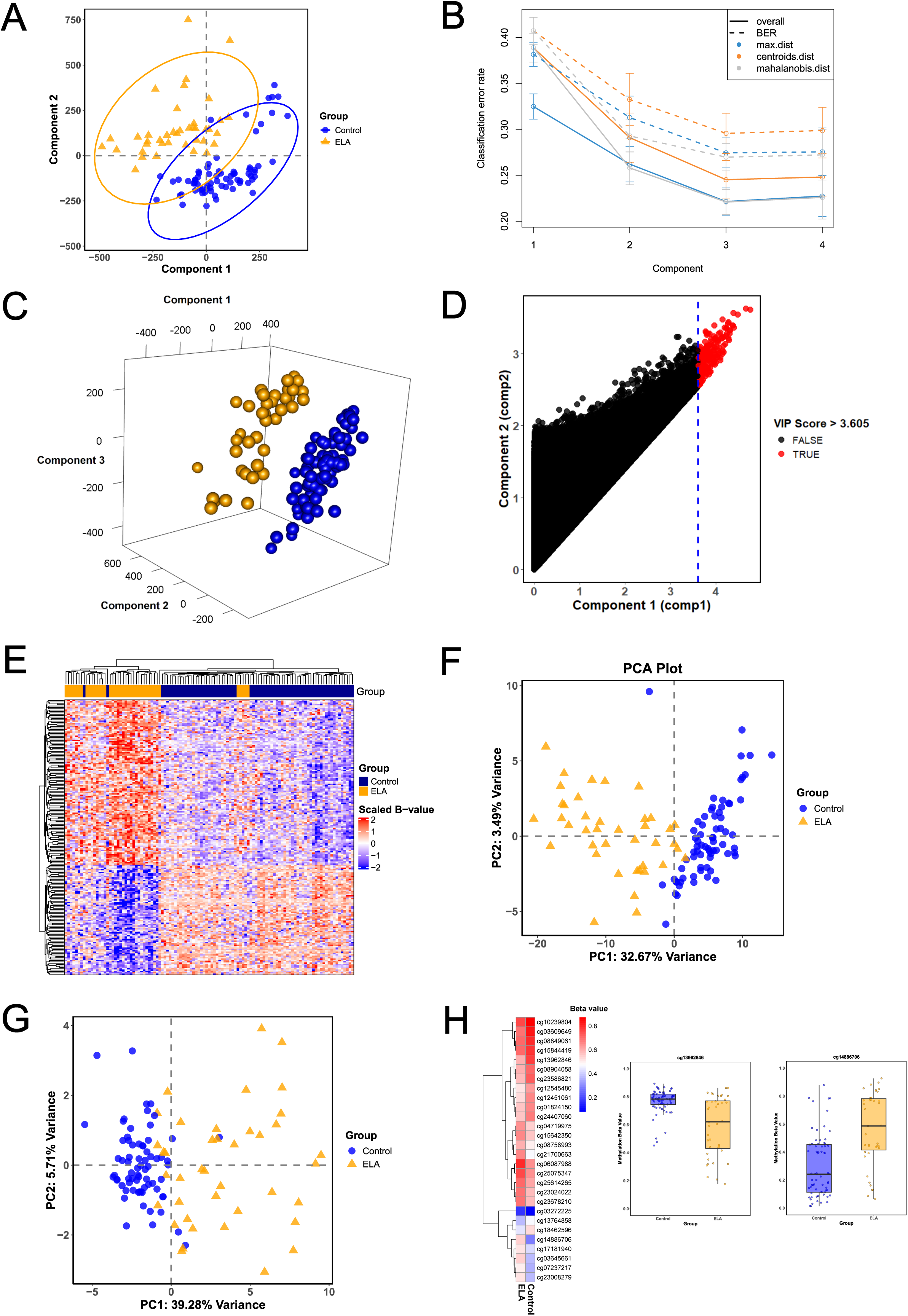
DNA methylation signature development using partial least squares discriminant analysis (PLS-DA) derived model. **(A)** Scatter plot shows PC1 nad PC2 after performing PLS-DA. **(B)** Graph shows model validation and the cumulative component classification-error rate. **(C)** 3-D graphical representation of the PLSDA model including 3 components from the discriminant analysis. **(D)** Scatter plot shows the distribution of variable importance projection (VIP) score. Blue dotted line represents the threshold set at 3.605. **(E)** Heatmap shows DNA methylation for CpG loci with FDR<0.05 and VIP>3.605. **(F)** PCA plot based on 200 CpG loci with FDR<0.05 and VIP>3.605. **(G)** PCA plot based on 28 CpG loci with FDR<0.05, effect size>10% and VIP>3.605. **(H)** Heatmap shows mean DNA methylation levels for the 28 most significant and biologically relevant CpGs making up the ELA epigenetic signature. Box plots represent selected probes cg08904058 and cg14886706. *p < 0.05, **p < 0.01, ***p< 0.001.

Variable Importance Projection (VIP) scores were calculated for each CpG locus to determine their contribution to the overall model. Using a stringent threshold of VIP > 3.605, we identified key CpG loci driving the classification (Figure 4D). By integrating the results from the EWAS (Figure 2A – B) and PLS-DA, we identified 200 CpG loci that were both differentially methylated (FDR < 0.05) and had a VIP score > 3.605 (Figure 4E – F). In addition, a PCA of the 200 CpGs showed a good separation between control and ELA groups with PC1 accounting for 32.67% of the total variance while PC2 explained 3.49% of the variance (Figure 4F). In order to refine our findings, we modified our selection criteria to CpG loci with FDR<0.05, effect size>10% and VIP>3.605. Using this stringent filter, we identified 28 CpG loci and performed another PCA. PC1 accounted for 39.28% of the total variance while PC2 explained 5.71% of the variance (Figure 4G). The analysis revealed a subset of 26 genes potentially associated with early-life adversity, including those previously implicated in stress response (JAK3, EPAS1, SENP1), neurodevelopment (ZEB2, SYT14, ITPR2), and mitochondrial metabolism (SLC8A1, EDC3). The potential role of these genes in mediating the long-term effects of ELA highlights key biological mechanisms underlying stress-related developmental outcomes.

### 3.5. KEGG, Reactome, and Gene Ontology Analysis reveal ELA-associated pathways

In order to identify biological processes affected by ELA we performed KEGG pathway analysis, Reactome signalling networks analysis, and Gene Ontology enrichment analysis. To strengthen our findings we employed an integrative approach that combined these analyses to identify key biological processes affected by ELA (Figure 5A). Pathways associated with endocrine and metabolic regulation such as insulin secretion, insulin resistance, cortisol synthesis, oxytocin signalling and PI3K-Akt signalling were significantly enriched (p<0.05). This aligns with previous studies suggesting that ELA increases the risk of metabolic disorders such as diabetes, obesity, and HPA axis dysregulation.

**Figure 5:**
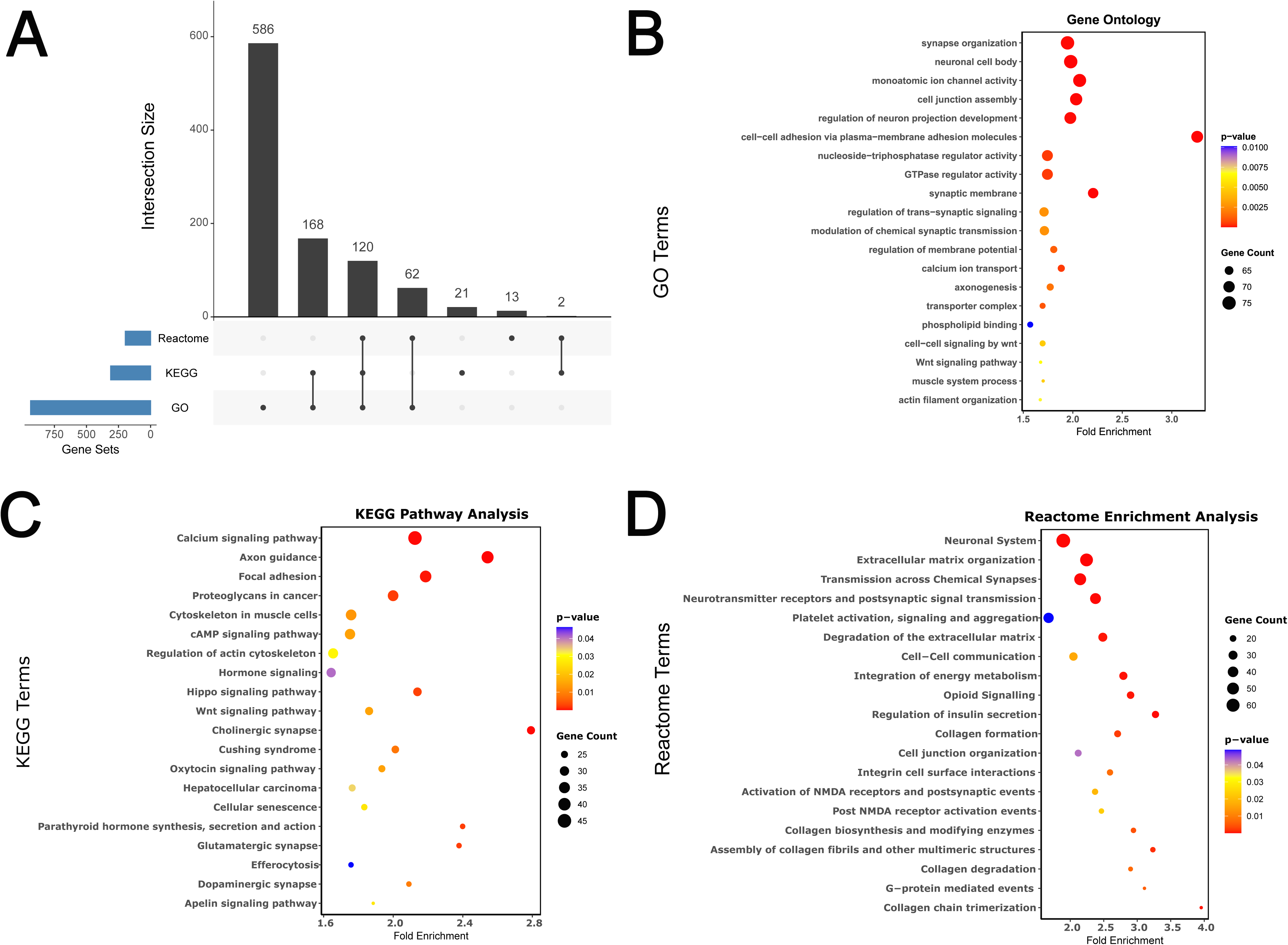
Integrated pathway analysis for differentially methylated CpG loci. **(A)** UpSet plot shows how genes identified in the different pathway analyses (Gene Ontology, KEGG and Reactome) intersect. **(B – D)** Plots shows the top 20 pathways for Gene Ontology, KEGG and Reactome, respectively.

Pathways related to immune response, including AGE-RAGE signalling in diabetic complications, leukocyte transendothelial migration, and cytokine-mediated signalling, were significantly enriched. GO analysis further highlighted processes such as regulation of immune response, inflammatory cytokine production, and modulation of chemotaxis (Figure 5B, Supplementary Table 4). These findings suggest that ELA-induced epigenetic modifications may be the underlying drivers of chronic low-grade inflammation and altered immune regulation. Consequently, these epigenetic aberrations increase the potential risk of autoimmune diseases and metabolic disorders later in life.

In addition to immune-related responses, pathways associated with neuronal function and synaptic regulation were also significantly enriched. KEGG analysis revealed strong associations with cholinergic, dopaminergic, glutamatergic, and GABAergic synapse pathways, while GO terms highlighted synapse organization, neurotransmitter transport, and regulation of trans-synaptic signalling (Figure 5A – B, Supplementary Table 5). These findings indicate that ELA may disrupt synaptic plasticity and cognitive function, contributing to an increased susceptibility to neuropsychiatric disorders such as depression and anxiety. In addition to neuronal pathways, calcium signaling and ion transport also emerged as key molecular processes affected by ELA. KEGG pathway analysis highlighted calcium ion transmembrane transport and voltage-gated calcium channel activity, while GO terms included membrane depolarization, regulation of membrane potential, and calcium ion import into the cytosol. These findings suggest that dysregulation of calcium homeostasis may contribute to altered neuronal excitability and synaptic transmission. Furthermore, dysregulation in calcium ion transport can contribute to cognitive deficits, heightened stress responses, and cardiovascular alterations. KEGG and Reactome analyses also showed enrichment in focal adhesion, extracellular matrix organization, and actin filament-based processes (Figure 5C, Supplementary Table 6). These findings suggest that ELA influences cellular architecture and structural integrity, potentially contributing to altered neuronal connectivity and tissue remodelling.

In line with our previous finding that ELA promotes senescence in NK cells, KEGG analysis revealed pathways related to cellular senescence (Figure 5C). Notably, our analysis also identified several novel pathways that have not been previously associated with ELA, namely, efferocytosis, Hippo signalling, glucuronosyltransferase activity, ceramide metabolism and GTPase-mediated signalling pathways.

### 3.6 DNA Methylation signature associates with ELA-related health outcomes in the EpiPath cohort

Having identified an epigenetic signature associated with ELA exposure, we examined how this signature associated with the phenotype observed in our cohort. We previously observed that 2 decades after exposure there were functional immune differences as well as overall health and health behaviour differences [18, 27, 28]. To investigate this relationship, we created a derivative dummy variable of the 28 CpG loci DNA methylation signature by extracting scores from the principal component analysis (PCA). We used PC1, which captured 39.28% of the total variance, as a summary measure of the DNA methylation signature.

Correlation analysis revealed a significant positive association with chronic diseases (r = 0.31, p < 0.05), implying that the epigenetic signature captured biologically relevant variation linked to long-term disease susceptibility (Figure 6A – B). PC1 also correlated with physical activity (r = 0.23, p<0.05; Figure 6C), but not with age, BMI, smoking and allergies. Additionally, we tested for correlation with cortisol levels and blood glucose levels but no significant associations were observed (Supplementary Figure 1). As a validation step, we examined the association between our epigenetic signature and CMV infection status, a critical confounder we had previously identified [16]. We found no significant relationship, confirming that the signature is not driven by CMV-related immune alterations.

**Figure 6:**
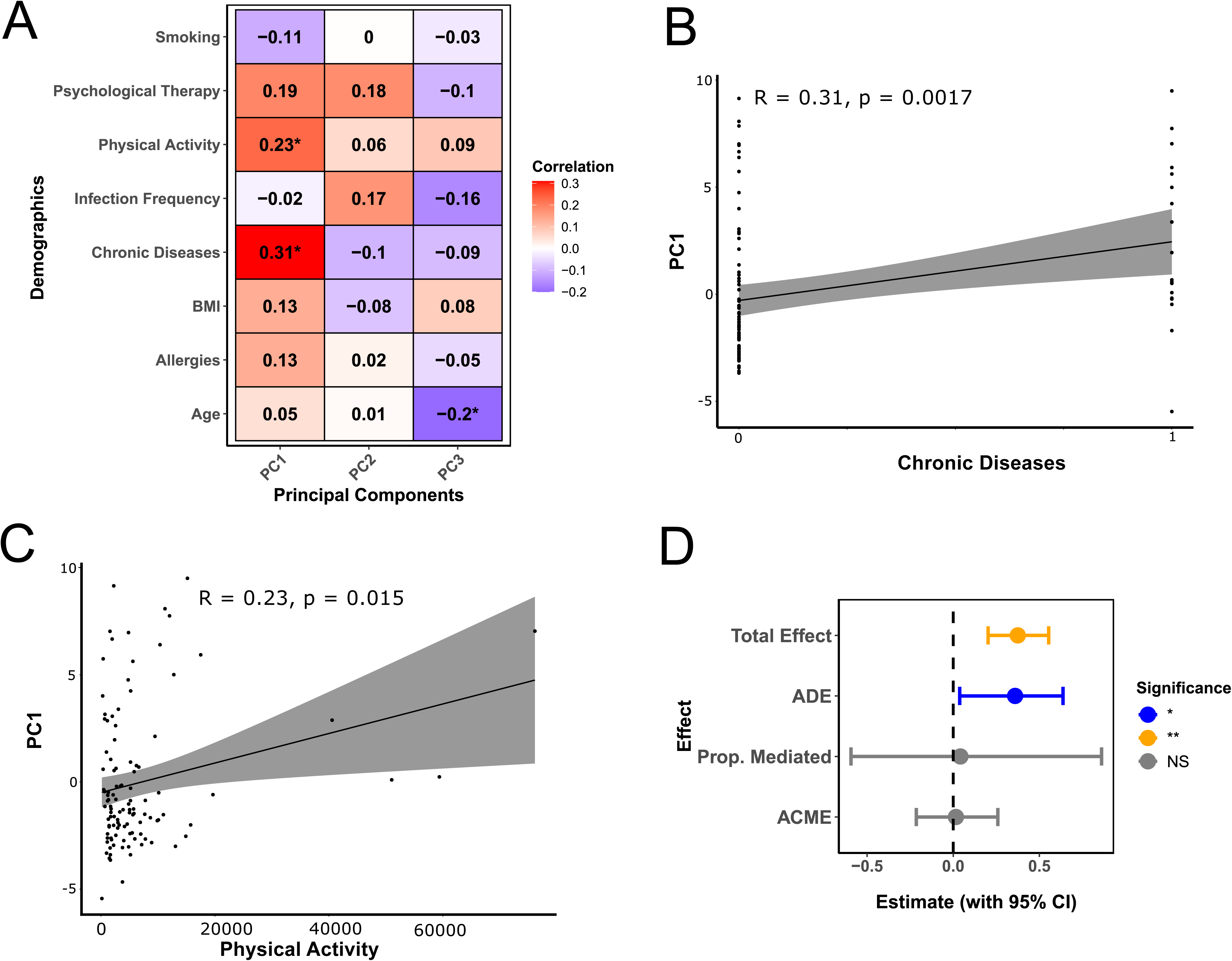
Correlation analysis and causal mediation analysis. **(A)** Heatmap shows correlation analysis results for PC1 against smoking, psychological therapy, physical activity, infection frequency, chronic diseases, BMI, allergies and age. Scatter plot shows correlation for PC1 vs **(B)** chronic diseases and **(C)** physical activity. **(D**) Forest plot shows the mediation analysis results with ELA-exposure as the treatment, PC1 (DNA methylation) as the mediator and chronic diseases as the health outcome. *p < 0.05, **p < 0.01, ***p< 0.001. Abbreviations; ACME, average causal mediation effect; ADE, average direct effect.

To determine the role of DNA methylation in linking ELA to chronic disease risk, we conducted a causal mediation analysis. The analysis revealed that the average causal mediation effect (ACME) was not significant (estimate = 0.0156, 95% CI = [-0.2143, 0.26], p = 0.892), indicating that DNA methylation did not significantly mediate the relationship between ELA and chronic disease risk (Figure 6D). However, the average direct effect (ADE) remained significant (estimate = 0.3591, 95% CI = [0.0375, 0.64], p = 0.018), as did the total effect (estimate = 0.3746, 95% CI = [0.2026, 0.55], p = 0.002), suggesting that ELA influences chronic disease risk through other mechanisms other than DNA methylation. The proportion mediated was estimated at 4.16% but was not statistically significant (p = 0.890), reinforcing the conclusion that while DNA methylation is associated with chronic diseases; it does not serve as a primary mediator.

## 4. Discussion

Our study underscores the enduring impact of ELA on long-term health, reinforcing Barker’s (1995) concept of sensitive periods [6, 29]. Unlike previous EWAS studies, our participants experienced brief adversity (median exposure 4.3 months [16]) before adoption, with assessments conducted 20-24 years later [16, 18, 28, 30, 31]. Our findings support the “three-hit” model, emphasizing the complex interplay between genetic predispositions and early environmental influences in shaping adult phenotypes [4, 5]. We identified widespread differential DNA methylation associated with ELA that persisted for two decades post-exposure. Significant changes were observed at both individual CpG loci and within differentially methylated regions (DMRs), particularly in promoter regions, suggesting potential effects on gene regulation. Principal component analysis (PCA) and partial least squares-discriminant analysis (PLS-DA) revealed distinct methylation patterns differentiating ELA and control groups, indicating a stable epigenetic signature maintained decades after exposure to ELA.

ELA encompasses stressors such as low socioeconomic status, parental separation, and institutionalization, all of which disrupt caregiver stability and impact lifelong health outcomes. Institutionalization, in particular, represents a strong psychosocial stressor [32–35] that has been linked to immune dysfunction and impaired natural killer (NK) cell functionality in adulthood [16, 18, 27]. Our study demonstrates that even a brief period of institutionalization (mean 4.3 months) in the early postnatal period leaves a lasting epigenetic mark. This finding aligns with previous research linking ELA severity to the age of adoption and length of institutionalization.

Pinpointing mechanisms driving low-grade inflammation in ELA-exposed individuals is crucial for understanding the persistence of the ELA phenotype. For instance, BDNF is a key player in modulating neuroinflammation[36], and its dysregulation following exposure to various adverse early life environments suggests that it is a versatile driver of the ELA phenotype. Consistent with previous findings an increase in *BDNF* methylation was observed in our study [37–39]. Differential methylation was also observed in genes associated with efferocytosis, a critical process for clearing apoptotic cells and maintaining immune homeostasis [40]. While chronic inflammation has been associated with ELA [8, 41], the role of impaired efferocytosis remains underexplored. Our findings suggest that disrupted efferocytosis may contribute to prolonged immune activation and increased autoimmune susceptibility. Additionally, differential methylation in glucuronosyltransferase family genes, including LARGE1, LARGE2, B3GAT1, EXTL1, and UGT1A subtypes [42] suggests a link between ELA and altered metabolism of stress hormones and neuroactive steroids. The dysregulation of glucuronidation may influence cortisol clearance, potentially explaining individual differences in stress resilience.

Epigenetic research into ELA continues to grow, particularly concerning exposure timing and duration [43]. Holuka et al. found persistent epigenetic modifications in adolescents born to mothers who experienced financial stress during pregnancy [44]. Similarly, Heijmans et al. demonstrated differential DNA methylation at *IGF2* loci in individuals exposed in utero to the Dutch Hunger Winter, suggesting long-term biological embedding of early adversity [45]. Our study identified differential methylation in CpG loci associated with *INS-IGF2* and *IGFBP3*, consistent with previous findings. Even though the timeframes of exposure are different, the duration of exposure may be a crucial determinant of affected genes and pathways. Furthermore, studies on prenatal stress, such as the 1998 Quebec Ice Storm, have linked DNA methylation changes to immune and metabolic pathways [46, 47]. Our findings parallel these observations, indicating that ELA consistently disrupts essential biological processes.

Pathway analysis revealed significant enrichment of genes involved in the oxytocin-signalling pathway, implicating epigenetic modifications in long-term social and emotional regulation [48]. Oxytocin is critical for social bonding, stress adaptation, and neuroendocrine function [9]. Methylation differences in *ZFP57*, a key regulator of maternal imprinting [49], suggest potential alterations in the expression of imprinted genes, such as SNRPN, which has been associated with autism and neurodevelopmental disorders [50]. These disruptions may contribute to long-term neurodevelopmental and metabolic consequences, given the role of imprinted genes in brain development and stress regulation. Furthermore, altered *ZFP57* methylation may affect the inheritance and maintenance of essential imprints, potentially influencing parent-of-origin gene expression patterns and increasing susceptibility to stress-related disorders across generations.

The Minnesota International Adoption Project showed that adolescence was a distinct period for recalibrating stress regulation through social and environmental influences, that we term the “Gunnar Window of Development” [51]. Unlike early childhood, where caregiver interactions primarily shape stress responses, adolescence represents a secondary critical period where peer relationships and social reorientation play a significant role in modifying long-term stress regulation [52]. While several studies suggest that recalibration of stress physiology is possible during this window [35, 53, 54], here, we found persistent DNA methylation signatures in individuals exposed to ELA pre-adolescence, and still present after this pubertal recalibration period. In our EpiPath cohort the HPA-axis reaction to a psychosocial stressor measured at the same time as the DNA was obtained for this study was dysregulated and had not been reprogrammed during adolescence [17]. This raises questions about sensitive periods, genomic imprinting, and the long-term programming of immune and neural systems. This agrees with our recent hypothesis that stable immunophenotypes in ELA individuals may be maintained through mechanisms such as DNA methylation in progenitor or stem cells [55], with the long-term generation of epignetically modified somatic cells.

Comparing our results with Naumova et al. [11, 14], we observed 16 overlapping differentially methylated genes despite differences in sampling timing. Naumova’s studies examined institutionalized children aged 8-35 months, whereas our cohort experienced adoption with blood sampling conducted in adolescence and young adulthood (18-33 years). The persistence of these differentially methylated genes across distinct cohorts suggests that ELA establishes enduring biological imprints. In support of this notion, a recent study in baboons, demonstrated that ELA leads to persistent DNA methylation changes that affect gene activity [56]. The current study extends this concept to humans, showing widespread DNA methylation changes that are associated with ELA. Our findings reinforce the importance of the type of exposure and timing of exposure [57] as key determinants of an individual’s health trajectory.

## 5. Conclusion

In conclusion, our study provides evidence that ELA leaves a lasting epigenetic imprint, as reflected in the DNA methylation signature that remains detectable decades after exposure. Our data shows that the epigenetic signature is not simply a reflection of immune activation but rather a biologically meaningful marker of ELA-related health trajectories. Defining “trajectory” in the context of adulthood diseases related to ELA remains complex. However, our study demonstrates that epigenetic marks persist beyond adolescence and are associated with chronic diseases. Despite individual variability, fewer than half of ELA participants developed chronic diseases by early adulthood, highlighting differences in health trajectories. Future research should refine trajectory classifications through data-driven approaches and explore the potential existence of a stress-exposure threshold, beyond which epigenetic reprogramming or recalibration becomes limited. Understanding biological mechanisms that drive the development of the ELA phenotype may help identify critical intervention windows and resilience factors in ELA-exposed individuals.

## 6. Conflict of Interest

The authors declare that the research was conducted in the absence of any commercial or financial relationships that could be construed as a potential conflict of interest.

## 7. Author Contributions

The EpiPath cohort was conceptualised by JDT and CPM. For this element of the study: conceptualization: AM and JDT; literature review: MB; data collection: SBM, AM, PG, MMCE, FADL; data curation and analysis: AM, MMCE, FADL, JLC; visualisation: AM; writing – original draft: AM and MB; scientific refinement – AM, MB, and JDT; manuscript review and editing: all authors. All authors read and approved the final version of the manuscript. Certain elements within the figures are from Biorender.com under the licence of JDT.

## 8. Funding

The Fonds National de Recherche Luxembourg funded the EpiPath Cohort (C12/BM/3985792 ‘EpiPath’); This study was funded by the Fonds National de Recherche Luxembourg grants FNR-CORE (C20/BM/14766620 “ImmunoTwin”) and the Ministry of Higher Education and Research of Luxembourg. MB was funded through (PRIDE23/18356118; XPose). The work of JDT on long term consequences of ELA was further funded by the FNR (C19/SC/13650569, “ALAC”; C16/BM/11342695, “MetCOEPs”; INTER/ANR/16/11568350 “MADAM”)

## Acknowledgments

The authors thank all the participants and their parents for taking part in this study. Special mention goes to Pauline Guebels for all the technical support in the laboratory.

## 9. Supplementary Material

Supplementary data for this article has been provided.

## 10. Data Availability

Data is available upon request to the authors.

## Notes

### Competing Interest Statement

The authors have declared no competing interest.

### Funding Statement

This study received funding from the Fonds National de Recherche Luxembourg (C12/BM/3985792, Epipath); Fonds National de Recherche Luxembourg grants FNR-CORE (C20/BM/14766620, ImmunoTwin) and the Ministry of Higher Education and Research of Luxembourg.

### Author Declarations

National Research Ethics Committee of Luxembourg and the University of Luxembourg Ethics Review Panel gave Luxembourg Institute of Health and the University of Luxembourg ethical approval for this work.

## References

1. Barker, D.J. and C. Osmond, Infant mortality, childhood nutrition, and ischaemic heart disease in England and Wales. Lancet, 1986. 1(8489): p. 1077–81.

2. Holuka, C., et al., Transgenerational impacts of early life adversity: from health determinants, implications to epigenetic consequences. Neurosci Biobehav Rev, 2024. 164: p. 105785.

3. Wadhwa, P.D., et al., Developmental origins of health and disease: brief history of the approach and current focus on epigenetic mechanisms. Semin Reprod Med, 2009. 27(5): p. 358–68.

4. Grova, N., et al., Epigenetic and Neurological Impairments Associated with Early Life Exposure to Persistent Organic Pollutants. Int J Genomics, 2019. 2019: p. 2085496.

5. Daskalakis, N.P., et al., The three-hit concept of vulnerability and resilience: toward understanding adaptation to early-life adversity outcome. Psychoneuroendocrinology, 2013. 38(9): p. 1858–73.

6. Barker, D.J., Fetal origins of coronary heart disease. BMJ, 1995. 311(6998): p. 171–4.

7. Hostinar, C.E., et al., Early-Life Socioeconomic Disadvantage and Metabolic Health Disparities. Psychosom Med, 2017. 79(5): p. 514–523.

8. Turecki, G., et al., Early life adversity, genomic plasticity, and psychopathology. Lancet Psychiatry, 2014. 1(6): p. 461–6.

9. Uvnas-Moberg, K., et al., The Yin and Yang of the oxytocin and stress systems: opposites, yet interdependent and intertwined determinants of lifelong health trajectories. Front Endocrinol (Lausanne), 2024. 15: p. 1272270.

10. Elwenspoek, M.M.C., et al., Glucocorticoid receptor signaling in leukocytes after early life adversity. Dev Psychopathol, 2020. 32(3): p. 853–863.

11. Naumova, O.Y., et al., Differential patterns of whole-genome DNA methylation in institutionalized children and children raised by their biological parents. Dev Psychopathol, 2012. 24(1): p. 143–55.

12. Wiik, K.L., et al., Behavioral and emotional symptoms of post-institutionalized children in middle childhood. J Child Psychol Psychiatry, 2011. 52(1): p. 56–63.

13. Tottenham, N., et al., Elevated amygdala response to faces following early deprivation. Dev Sci, 2011. 14(2): p. 190–204.

14. Naumova, O.Y., et al., Effects of early social deprivation on epigenetic statuses and adaptive behavior of young children: A study based on a cohort of institutionalized infants and toddlers. PLoS One, 2019. 14(3): p. e0214285.

15. Gunnar, M.R., et al., Pubertal stress recalibration reverses the effects of early life stress in postinstitutionalized children. Proc Natl Acad Sci U S A, 2019. 116(48): p. 23984–23988.

16. Elwenspoek, M.M.C., et al., T Cell Immunosenescence after Early Life Adversity: Association with Cytomegalovirus Infection. Front Immunol, 2017. 8: p. 1263.

17. Hengesch, X., et al., Blunted endocrine response to a combined physical-cognitive stressor in adults with early life adversity. Child Abuse Negl, 2018. 85(1873-7757 (Electronic)): p. 137–144.

18. Elwenspoek, M.M.C., et al., Proinflammatory T Cell Status Associated with Early Life Adversity. J Immunol, 2017. 199(12): p. 4046–4055.

19. Elwenspoek, M.M.C., Phenotype and Mechanisms of Altered Immune Functions induced by Early Life Adversity Phänotyp und Mechanismen veränderter Immunfunktionen, induziert durch traumatische Erfahrungen in der frühen Kindheit. 2018.

20. Zhou, W., et al., SeSAMe: reducing artifactual detection of DNA methylation by Infinium BeadChips in genomic deletions. Nucleic Acids Res, 2018. 46(20): p. e123.

21. Leek, J.T., et al., The sva package for removing batch effects and other unwanted variation in high-throughput experiments. Bioinformatics, 2012. 28(6): p. 882–3.

22. Wickham, H., ggplot2: Elegant Graphics for Data Analysis. 2016, New York: Springer-Verlag.

23. Wilke, C., cowplot: Streamlined Plot Theme and Plot Annotations for ‘ggplot2’. 2024.

24. Wei, T. and V. Simko, R package ‘corrplot’: Visualization of a Correlation Matrix. 2024.

25. Gu, Z., Complex heatmap visualization. Imeta, 2022. 1(3): p. e43.

26. Kolde, R., pheatmap: Pretty Heatmaps. 2019.

27. Fernandes, S.B., et al., Unbiased Screening Identifies Functional Differences in NK Cells After Early Life Psychosocial Stress. Front Immunol, 2021. 12: p. 674532.

28. Hengesch, X., et al., Blunted endocrine response to a combined physical-cognitive stressor in adults with early life adversity. Child Abuse Negl, 2018. 85: p. 137–144.

29. Lussier, A.A., et al., Sensitive Periods for the Effect of Childhood Adversity on DNA Methylation: Updated Results From a Prospective, Longitudinal Study. Biol Psychiatry Glob Open Sci, 2023. 3(3): p. 567–571.

30. Charalambous, E.G., et al., Early-Life Adversity Leaves Its Imprint on the Oral Microbiome for More Than 20 Years and Is Associated with Long-Term Immune Changes. Int J Mol Sci, 2021. 22(23).

31. Charalambous, E.G., et al., The oral microbiome is associated with HPA axis response to a psychosocial stressor. Sci Rep, 2024. 14(1): p. 15841.

32. van, I.M.H., et al., Children in Institutional Care: Delayed Development and Resilience. Monogr Soc Res Child Dev, 2011. 76(4): p. 8–30.

33. Gunnar, M.R., J. Bruce, and H.D. Grotevant, International adoption of institutionally reared children: research and policy. Dev Psychopathol, 2000. 12(4): p. 677–93.

34. Gunnar, M.R., M.H. van Dulmen, and T. International Adoption Project, Behavior problems in postinstitutionalized internationally adopted children. Dev Psychopathol, 2007. 19(1): p. 129–48.

35. Gunnar, M.R., et al., Moderate versus severe early life stress: associations with stress reactivity and regulation in 10-12-year-old children. Psychoneuroendocrinology, 2009. 34(1): p. 62–75.

36. Charlton, T., et al., Brain-derived neurotrophic factor (BDNF) has direct anti-inflammatory effects on microglia. (1662-5102 (Print)).

37. Duffy, H.B.D. and T.L. Roth, Increases in Bdnf DNA Methylation in the Prefrontal Cortex Following Aversive Caregiving Are Reflected in Blood Tissue. Front Hum Neurosci, 2020. 14: p. 594244.

38. Kundakovic, M., et al., DNA methylation of BDNF as a biomarker of early-life adversity. Proc Natl Acad Sci U S A, 2015. 112(22): p. 6807–13.

39. Rivera, L.M., et al., Prenatal exposure to genocide and subsequent adverse childhood events are associated with DNA methylation of SLC6A4, BDNF, and PRDM8 in early adulthood in Rwanda. (2045-2322 (Electronic)).

40. Cai, W., et al., STAT6/Arg1 promotes microglia/macrophage efferocytosis and inflammation resolution in stroke mice. JCI Insight, 2019. 4(20).

41. McGowan, P.O., et al., Epigenetic regulation of the glucocorticoid receptor in human brain associates with childhood abuse. Nat Neurosci, 2009. 12(3): p. 342–8.

42. Bock, K.W., Roles of human UDP-glucuronosyltransferases in clearance and homeostasis of endogenous substrates, and functional implications. Biochem Pharmacol, 2015. 96(2): p. 77–82.

43. Lussier, A.A., et al., Association between the timing of childhood adversity and epigenetic patterns across childhood and adolescence: findings from the Avon Longitudinal Study of Parents and Children (ALSPAC) prospective cohort. Lancet Child Adolesc Health, 2023. 7(8): p. 532–543.

44. Holuka, C., et al., Developmental epigenomic effects of maternal financial problems. Dev Psychopathol, 2024: p. 1–14.

45. Heijmans, B.T., et al., Persistent epigenetic differences associated with prenatal exposure to famine in humans. Proc Natl Acad Sci U S A, 2008. 105(44): p. 17046–9.

46. Provencal, N., et al., Glucocorticoid exposure during hippocampal neurogenesis primes future stress response by inducing changes in DNA methylation. Proc Natl Acad Sci U S A, 2020. 117(38): p. 23280–23285.

47. Cao-Lei, L., et al., DNA methylation mediates the impact of exposure to prenatal maternal stress on BMI and central adiposity in children at age 13(1/2) years: Project Ice Storm. Epigenetics, 2015. 10(8): p. 749–61.

48. Uvnas Moberg, K., et al., Editorial: Sensory Stimulation and Oxytocin: Their Roles in Social Interaction and Health Promotion. Front Psychol, 2022. 13: p. 929741.

49. Li, X., et al., A maternal-zygotic effect gene, Zfp57, maintains both maternal and paternal imprints. Dev Cell, 2008. 15(4): p. 547–57.

50. Li, H., et al., The autism-related gene SNRPN regulates cortical and spine development via controlling nuclear receptor Nr4a1. Sci Rep, 2016. 6: p. 29878.

51. Gunnar, M.R. and M.A. Howland, Calibration and recalibration of stress response systems across development: Implications for mental and physical health. Adv Child Dev Behav, 2022. 63: p. 35–69.

52. Silvers, J.A., Adolescence as a pivotal period for emotion regulation development. Curr Opin Psychol, 2022. 44: p. 258–263.

53. Gunnar, M.R., et al., Developmental changes in hypothalamus-pituitary-adrenal activity over the transition to adolescence: normative changes and associations with puberty. Dev Psychopathol, 2009. 21(1): p. 69–85.

54. Perry, N.B., et al., Cortisol Reactivity and Socially Anxious Behavior in Previously Institutionalized Youth. Res Child Adolesc Psychopathol, 2022. 50(3): p. 375–385.

55. Mposhi, A. and J.D. Turner, How can early life adversity still exert an effect decades later? A question of timing, tissues and mechanisms. Front Immunol, 2023. 14: p. 1215544.

56. Anderson, J.A., et al., DNA methylation signatures of early-life adversity are exposure-dependent in wild baboons. Proc Natl Acad Sci U S A, 2024. 121(11): p. e2309469121.

57. Suglia, S.F., et al., Timing, duration, and differential susceptibility to early life adversities and cardiovascular disease risk across the lifespan: Implications for future research. (1096-0260 (Electronic)).

