## Supplementary figures and images for "Epigenetic Signatures Reveal Biological Embedding of the Early-Life Environment Two Decades after Exposure to Adversity"

### Supplementary Figure 1.pdf

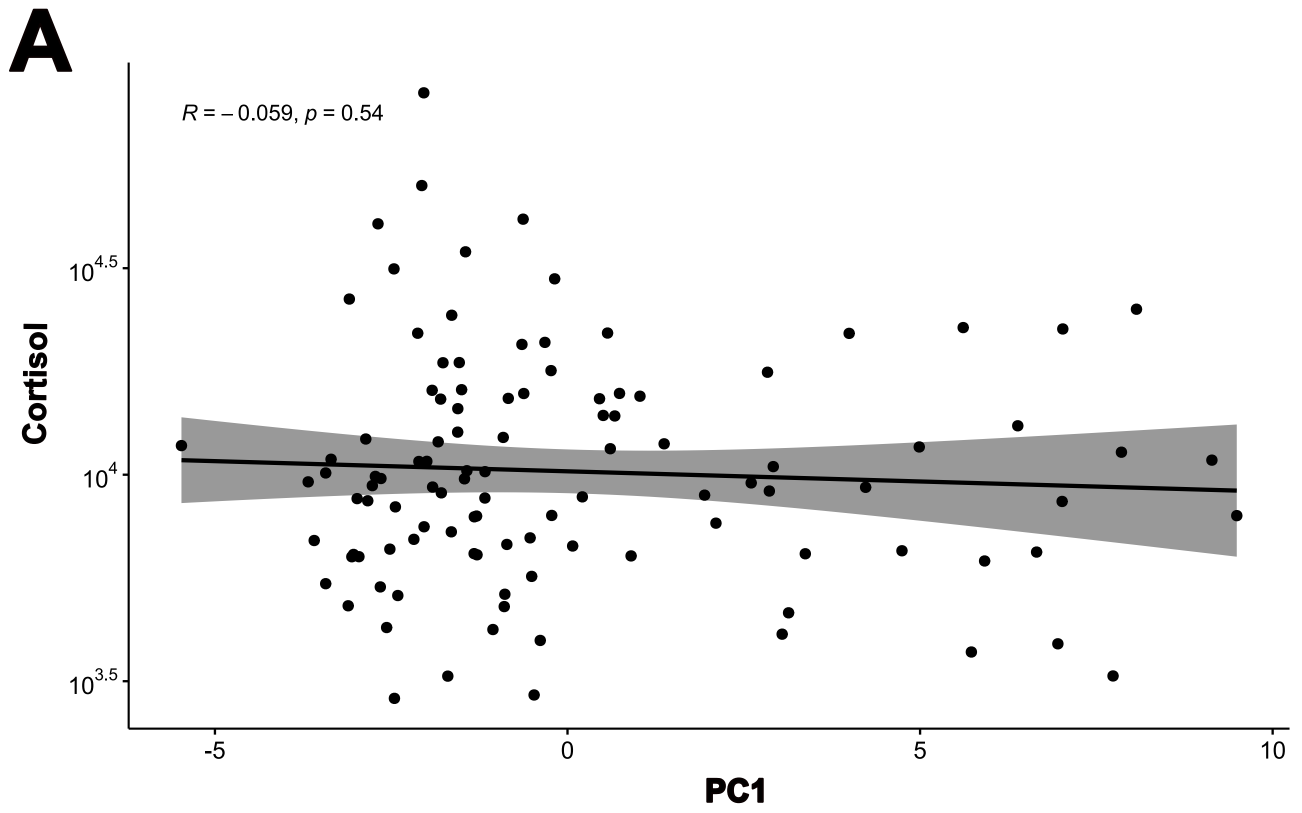

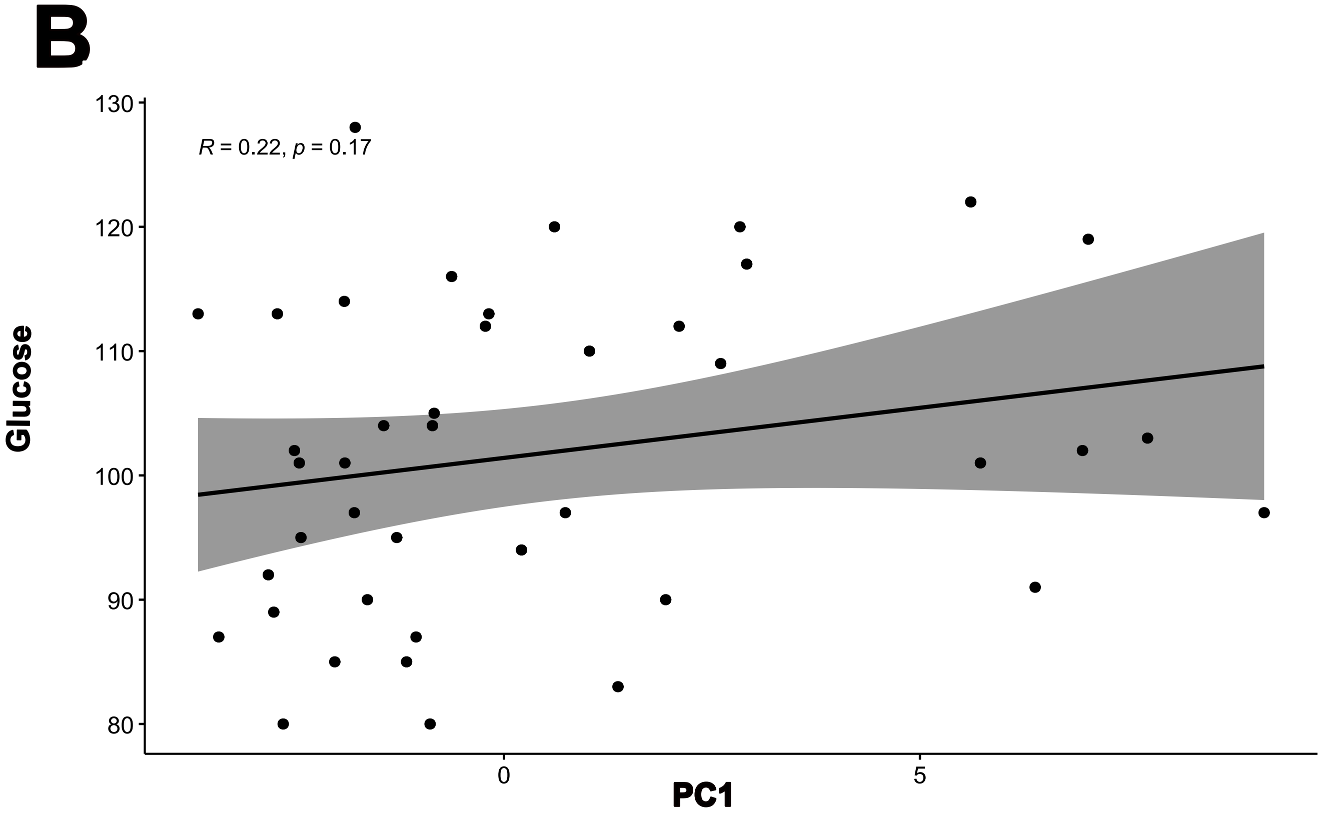
